# The detection of SARS-CoV-2 in autolysed samples from an exhumed decomposed body: Implications to virus survival, genome stability and spatial distribution in tissues

**DOI:** 10.1101/2021.02.16.21251805

**Authors:** Mahadeshwara Prasad, Somanna Ajjamada Nachappa, Niveditha Anand, Deepika Udayawara Rudresh, Yashika Singh, Surabhi P. Gangani, Forum K. Bhansali, Basista Rabina Sharma, Deep Nithun Senathipathi, Shashidhar H. Byrappa, Prakash M. Halami, Ravindra P. Veeranna, CSIR, CFTRI

## Abstract

Here we report for the first time the SARS-CoV-2 detection in autolysed samples from an exhumed decomposed body post-thirty six days after death. Both naso-oropharyngeal swabs and visceral samples from the lung, intestine, liver, and kidney were collected from the body exhumed post-fifteen days after burial, stored in viral transport medium and in saturated salt solution respectively. Naso-oropharyngeal swabs showed the presence of the SARS-CoV-2 genome as identified by the amplification of viral E, N, RdRP, or ORF1ab genes by RT-PCR. Subsequent examination of tissues reveal the detection of the virus genome in the intestine and liver, while no detection in the kidney and lung. These results signify the genome stability and implicate the virus survival in decomposed swab samples and in tissues and thereafter in storage solution. Further results also indicate spatial distribution of the virus in tissues during the early stage of infection in the subject with no respiratory distress. Considering the presence of cool, humid, and moist location of the exhumation, the presence of virus genome might also indicate that SARS-CoV-2 can persist for more than seven days on the surface of dead bodies similar to the Ebola virus, confirming that transmission from deceased subjects is possible for an extended period after death. These results further reaffirm the robustness of the RT-PCR aiding in the detection of viruses or their genome in decomposed samples when other methods of detection could not be useful.

## Background

Severe Acute Respiratory Syndrome-Corona Virus (SARS-CoV)-2 causes coronavirus disease (COVID)-19 characterized by fever, dry cough, body pain, loss of smell, taste, respiratory distress, multi-organ failure, and death[1].The case fatality rate (CFR) of the disease varies between 2%-13%, majority of people aged over 60 years and people with impaired immune function, and those with underlying medical conditions including diabetes, cardiovascular diseases, etc., or are most affected [2],[3],[4]. So far, 90 M people were found positive, 50 M people recovered, while 2 M have died globally[5]. The incubation period of the disease varies between 2-14 days[6]. The transmission mode includes surface contact of aerosol droplets from infected persons, followed by touching the nose, eyes, and mouth. Evidence also points towards vertical transmission to new-borns and faecal transmission[7],[8]. The efforts to mitigate the spread of the virus infection depend on high testing-to-case ratio, contact tracing, quarantine or isolation, and treatment strategies of positives[9], vaccination and following the COVID-19 appropriate behaviour such as frequent hand sanitization, face masking, social distancing[10].

Detection of viral RNA by the real-time reverse transcription-polymerase chain reaction (RT-PCR) remains the gold-standard technique of confirmation of COVID-19. The RT-PCR method is very sensitive and specific in detecting the SARS-CoV-2 virus genome[11]. The interpretation of results depends on the accuracy of the test and the pre-test probability or estimated risk of disease before testing[12],[13]. Since RT-PCR detection of the virus is based on the viral genome amplification, the presence of the viral RNA alone, irrespective of the virus’s viability or infectivity, could show positive[13]. There are reports that recovered people without clinical disease showing positive by RT-PCR due to traces of fragmented RNA in the naso-oropharyngeal region[14]. Further, there are reports that suggest differential sensitivity rate in the detection of the virus for specimens obtained from different sites indicating potential diversity in the distribution of virus in different mucosal surfaces and parts of the tissues such as the lung, intestine, liver, kidney etc.[15]. The dynamics of virus shedding, viral load from other sites, and time of infection have been thought to account for this variation in detection of the virus, for example, the virus being present in deeper respiratory specimens with the advanced disease[16]. However, more studies are required to understand the significance of sampling at different sites in the context of temporal and spatial distribution of virus in the body. A negative PCR result needs to be interpreted in the context of this variability in viral shedding. A negative PCR assay could be due to a true negative that people have not been infected with the SARS-CoV-2 or due to sampling error, sampling timing, the viral load, viral shedding, and the virus’s presence in deeper respiratory tissues as noted under the advanced stage[17],[18]. Current practice is to repeat further naso-oropharyngeal swabs or, if possible, to take deep respiratory samples if the first naso-oropharyngeal swab is negative. When a patient is suspected of having COVID-19, the current recommendation is to take two swabs each from upper and lower respiratory tracts[19].

The degree to which live viruses can survive in various environments and dead human tissue has been the subject of intense debate since the beginning of this pandemic[20],[21]. This critical piece of information can substantially impact a broad spectrum of areas, from the safe handling of laboratory specimens to disease mitigation procedures and the disposal of the dead body[19]. The RNA has been recovered from the 1918 influenza epidemic using pathology museum samples and lung tissue samples obtained from exhumed bodies from a mass grave in Alaska as late as 1997, though its viability is debatable[22]. Further, the oldest viral genome extracted and sequenced belongs to the hepatitis B virus[23]. To date, there has been no published data on the persistence of SARS-CoV-2 or its genomic RNA in the naso-oropharyngeal swabs, and in various tissues such as the lung, intestine, liver and kidney from the exhumed decomposed body, and we believe this is the first report to illustrate that SARS-CoV-2 was detected in decomposed naso-oropharyngeal swabs and tissue samples. This novel finding signifies the fact that SARS-CoV-2 can be detected in decomposed samples and swabbing upper respiratory mucosa is sufficient for obtaining samples for diagnosis. In addition, the findings also implicate virus survival and genome stability, and spatial distribution of virus in tissues in early infection. Furthermore, results also suggests that SARS-CoV-2 similar to other enveloped viruses such as Ebola virus[24] can persist for more than seven days on the surface of dead bodies and in body tissues, confirming that transmission from deceased subjects is possible for an extended period after death. These results further reaffirm the robustness of the RT-PCR aiding in the detection of viruses in exhumed, decomposed samples when other methods of detection could not be useful.

### Case presentation and qualitative multiplex one-step RT-PCR

Medical history reveals of a subject with age in 40’s, had fever, cough, body pain with no other apparent health issues was earlier tested COVID-19 negative by rapid antigen test (RAT) and positive by RT-PCR. Two days post-detection, the subject died under mysterious circumstances and the body was buried in an eight feet deep pit under a tree surrounded by a cool, humid location with paddy fields around. To ascertain the cause, mode and manner of the death, fifteen days post-burial; the body was exhumed and post-mortem examination was conducted as per the law by the forensic medicine specialist. The ethical approval for the testing of samples was obtained from the Mysore Medical College & Research Institute (MMC&RI), Mysuru (MMCEC24/20). The body showed all features of decomposition. The relaxed joints, flabby muscles, fallen and easily pluckable hairs, protruded eyeballs, swollen tongue, abdomen, and genitals. The whole body was moist and foul-smelling with flies flying around it. The superficial skin layer of the body was peeled entirely and destroyed. Internal examination of the body revealed that the brain was liquified, foul-smelling, lungs, liver, and spleen, stomach, small and large intestine were filled with gas. The mucosa of the larynx and trachea was disintegrated with intact cartilage. The oropharyngeal and nasopharyngeal swab samples were collected in viral transport medium (VTM) and tissues such as lung, intestine, liver, kidney were collected in saturated solution of sodium chloride and stored in −80°C freezer. Post-twenty one days after collection of samples, following completion of legal procedures, samples were sent for the detection of SARS-CoV-2.

The RNA was extracted from naso-oropharyngeal samples and from tissues using RNA spin columns manually (HiMedia, Mumbai) as well as using automated RNA extractor (Genetix Biotech Asia Pvt. Ltd., New Delhi) as per the protocol described by each kit. The extracted RNA was reconstituted in RNase free water and stored in −80°C deep freezer until the RT-PCR assay was set up. The real-time one-step multiplex RT-PCR was done using TaqMan probes designed to target genes coding for envelope (E), nucleoprotein (N), open reading frame 1ab (ORF1ab) and or RNA dependent RNA polymerase (RdRP) with RNase P or actin as internal control as specified by each kit [(Genes2Me (Genes2Me Pvt. Ltd., Gurugram), Q-Line molecular (POCT Services, New Delhi), and Meril (Meril Diagnostics Pvt. Ltd., Vapi)]. All these kits have been approved by the Indian Council of Medical Research (ICMR, New Delhi) for detection of SARS-CoV-2 in clinical samples. The RNase free water as template for negative controls, extracted RNA as template for the test, and respective template provided with the kit for positive controls were included for the detection. The PCR cycle conditions were set as determined by each kit. The total number of cycles fixed to forty. The cycle threshold (CT) values and the amplification curve were recorded. Mean CT values were used for the interpretation of results.

## Results and discussion

### Naso-oropharyngeal swabs showed positive for SARS-CoV-2 by RT-PCR

Naso-oropharyngeal swab samples were processed and the extracted RNA examined for the presence of SARS-CoV-2 by RT-PCR. Results show that mean CT values for E, N, RdRP, and RNase P genes were 27.1, 25.3, 25.8, and 34.0, respectively using the Genes2Me kit. The viral gene amplification was further examined using the other kit (Q-line), that adopts the dual-target gene design, which targets specific conserved sequence encoding the ORF1ab gene and the N gene. The mean CT values of N, ORF1ab, and that of the internal control were 29.4, 28.8, 30.4, respectively. Furthermore, results were additionally verified by using another kit (Meril) which also utilizes dual gene amplification for detection. The mean CT values observed were 31 for N gene, 31.5 for ORF1ab gene and 34.5 for the RNase P internal control gene. Fig. 1 shows the linear amplification plot of the above genes using all three kits for the negative control (left panel), test (mid panel), and that of the positive control (right panel). These results confirm the presence of the SARS-CoV-2 RNA in the naso-oropharyngeal samples from the exhumed decomposed body post-thirty six days after death.

**Fig. 1:**
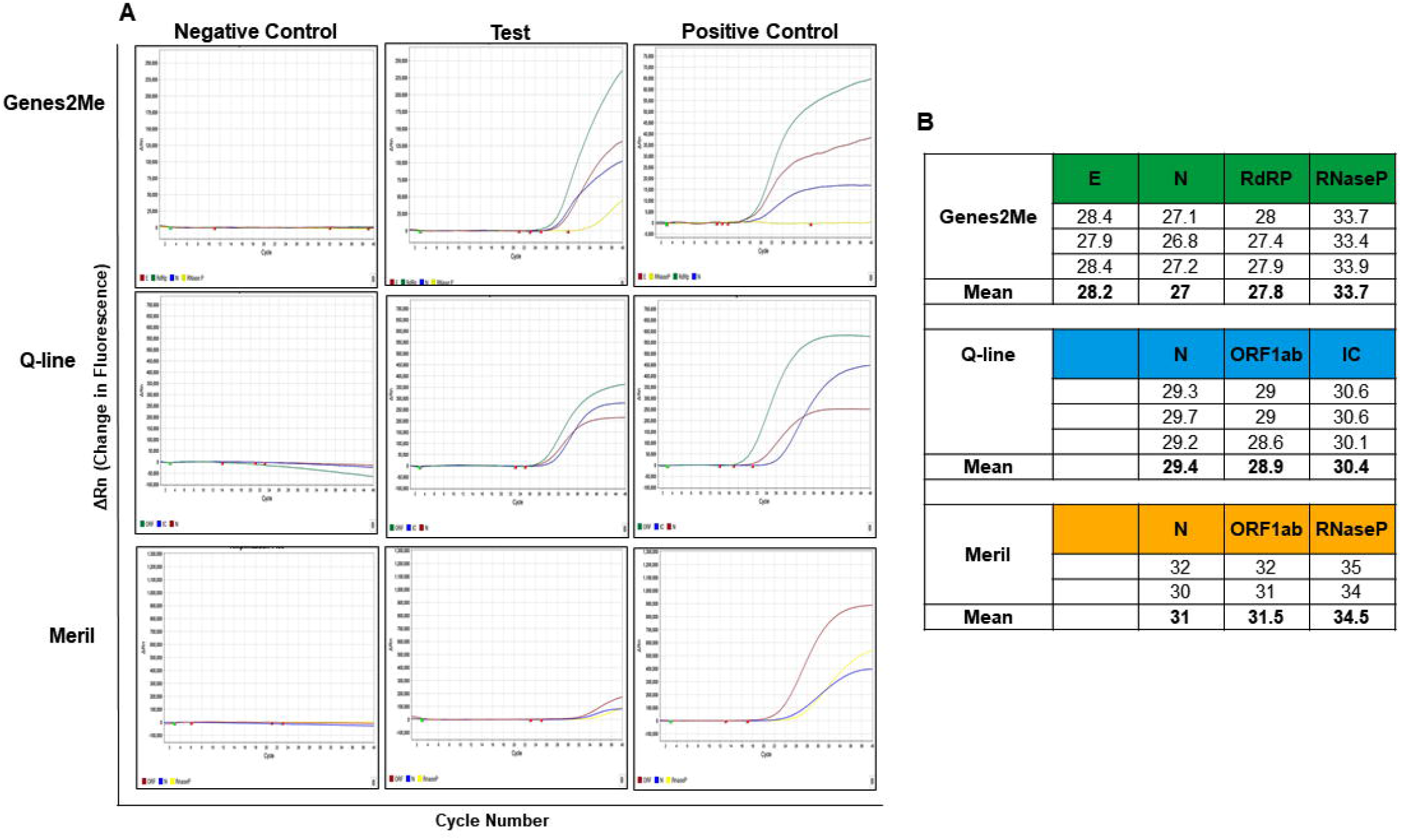
**A:** The linear plot of amplification curves showing change in fluorescence (ΔRn) Vs. cycle number following RT-PCR of naso-oropharyngeal swab samples using three different RT-PCR kits. **B:** Table showing the cycle threshold (CT) of samples in triplicate set.

### Intestine, liver tissues showed positive while the kidney and lung tissues showed negative for SARS-CoV-2

Microscopy examination of the lung and kidney revealed autolytic changes (data not shown). Therefore, to examine if SARS-CoV-2 RNA could be detected in tissues, the lung, intestine, liver and kidney tissues were processed and examined by RT-PCR. As shown in the figure 2, the intestine, liver showed amplification however, the kidney and the lung did not show the amplification of SARS-CoV-2 specific E, N, RdRP or ORF1ab genes as verified using all three kits. The mean CT values of the above genes are shown in the table 1.

**Table 1:** Table showing the cycle threshold (CT) values of tissue samples in triplicate set.

| Kit | Lung |  |  |  | Intestine |  |  |  | Liver |  |  |  | Kidney |  |  |  |
| --- | --- | --- | --- | --- | --- | --- | --- | --- | --- | --- | --- | --- | --- | --- | --- | --- |
| Genes2Me | E | N | RdRP | RNaseP | E | N | RdRP | RNaseP | E | N | RdRP | RNaseP | E | N | RdRP | RNaseP |
|  | - | - | - | 21 | 24.2 | 25.1 | 22.9 | 22.8 | 24.6 | 27.8 | 29.7 | 19.5 | - | - | - | 20.8 |
|  | - | - | - | 22.5 | 24.8 | 23.1 | 23.8 | 23.7 | 26.1 | - | 33.9 | 20.4 | - | - | - | 21.9 |
|  | - | - | - | 23.2 | 25 | 24.5 | 24.3 | 25 | 20.7 | 20.9 | 35.8 | 19.6 | - | - | - | 21.9 |
| Mean |  |  |  | 22.2 | 24.7 | 24.2 | 23.7 | 23.9 | 23.8 | 24.4 | 33.1 | 19.8 |  |  |  | 21.5 |

| Q-line |  | N | ORF1ab | IC | - | N | ORF1ab | IC |  | N | ORF1ab | IC |  | N | ORF1ab | IC |
| --- | --- | --- | --- | --- | --- | --- | --- | --- | --- | --- | --- | --- | --- | --- | --- | --- |
|  |  | - | - | 36.7 | - | 29 | 28.2 | 23.1 |  | 24 | 26.9 | 19.2 |  | - | - | 21.2 |
|  |  | - | - | 36.9 | - | 23.8 | 27.6 | 21 |  | 16.2 | 20.5 | 18.6 |  | - | - | 20 |
|  |  | - | - | 37 | - | 25.7 | 27.7 | 24.6 |  | 11.5 | 18.3 | 17.6 |  | - | - | 20.6 |
| Mean |  |  |  | 36.9 |  | 26.2 | 27.8 | 22.9 |  | 17.2 | 21.9 | 18.5 |  |  |  | 20.6 |

Table. 1
| Meril |  | N | ORF1ab | RNaseP |  | N | ORF1ab | RNaseP |  | N | ORF1ab | RNase P |  | N | ORF1ab | RNaseP |
| --- | --- | --- | --- | --- | --- | --- | --- | --- | --- | --- | --- | --- | --- | --- | --- | --- |
|  |  | - | - | 33.8 |  | 27.4 | 28 | 22.7 |  | 25.4 | 27.2 | 18.6 |  | - | - | 20.7 |
|  |  | - | - | 36.7 |  | 27.3 | 28.4 | 21 |  | 19.2 | 20.6 | 18.2 |  | - | - | 20.8 |
|  |  | - | - | 35.8 |  | 27.4 | 27.4 | 25 |  | 18.3 | 23.5 | 18.3 |  | - | - | 20.9 |
| Mean |  |  |  | 35.4 |  | 27.4 | 27.9 | 22.9 |  | 21 | 23.8 | 18.4 |  |  |  | 20.8 |

**Fig. 2:**
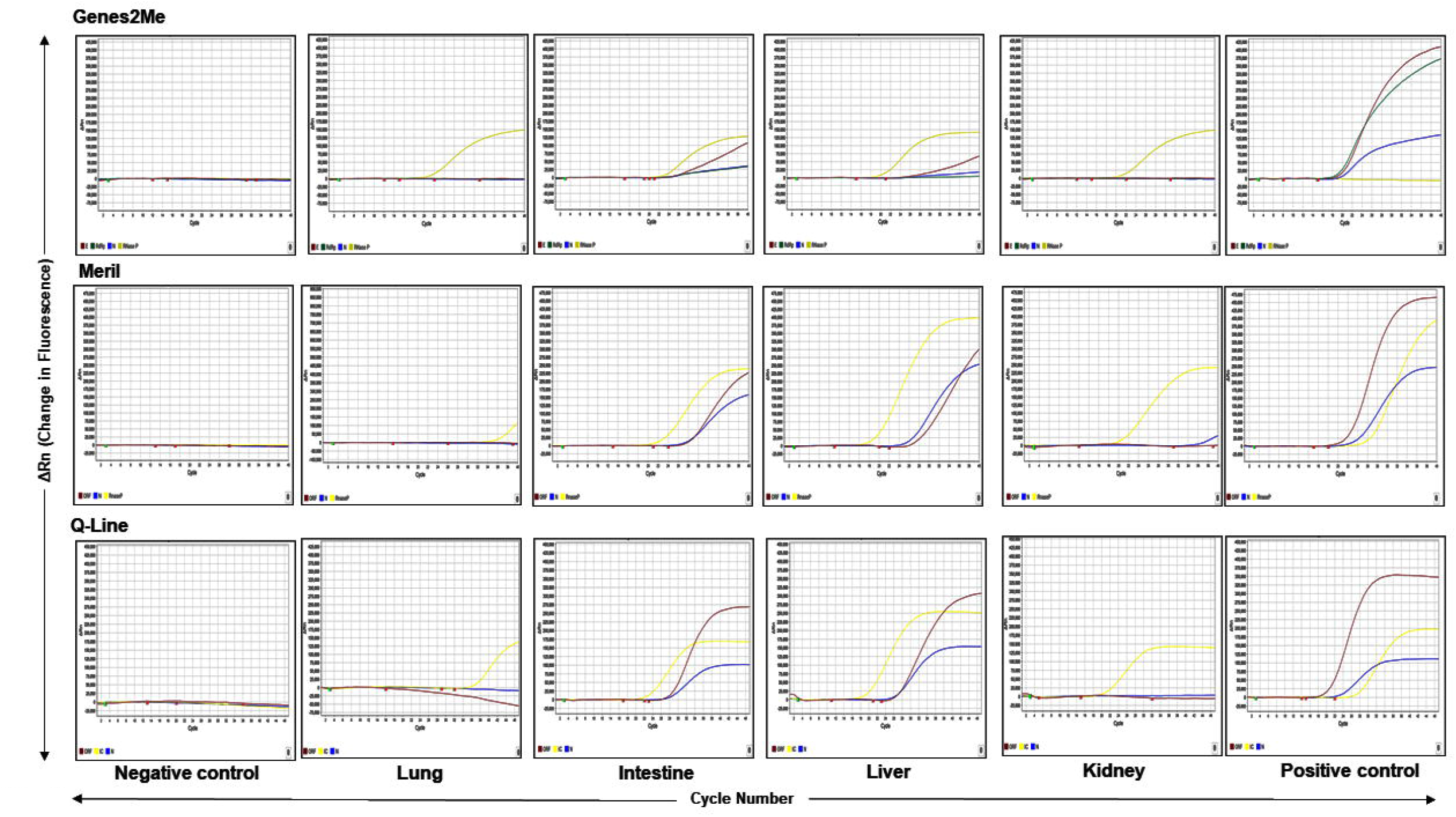
The linear plot of amplification curves showing change in fluorescence (ΔRn) Vs. cycle number following RT-PCR of autolysed tissue samples lung, intestine, liver and kidney) using three different RT-PCR kits.

These results indicate stability of the SARS-CoV-2 genome as well as virus survival in decomposed samples. Further, it also suggests temporal and spatial distribution of the virus in tissues. Additionally, it also confirms the utility of the PCR for the detection of viruses in samples from decomposed tissues where other techniques are not useful. Morkotter et al. (2015)[25] utilized RT-PCR assay to diagnose rabies infection in exhumed dog carcasses when other methods were unsuccessful because of the state of decomposition of the brain material. Further, they observed that the RT-PCR method was useful for forensic examination of decomposed tissue and to obtain the epidemiological information which otherwise would not have known with other conventional methods. In this context, RT-PCR assay was useful for the detection of SARS-CoV-2 in these decomposed samples while other methods including histopathology, and high-resolution computed tomography (HRCT), blood examination etc., have limitations. However, RT-PCR, being a very sensitive and genome-based technique can show positive with the mere presence of traces of fragmented genome, which is observed in samples of asymptomatic and recovered subjects[26]. Additionally, findings from this assay might not reveal if the virus was viable or dead in the decomposed tissue but demonstrate the stability of the SARS-CoV-2 genome in the decomposed tissue. Though there is no data yet as it relates to the detection of SARS-CoV-2 by RT-PCR in dead and decomposed tissues from the exhumed body, based on information from other viruses, typically, if an infectious virus is detected for days to weeks after the death, the genome of the virus can be detected for months to years.

The ability of the SARS-CoV-2 to survive in different environmental conditions like soil, varying humidity and temperature is not adequately documented. Further, there are no specific studies relating to the stability of SARS-CoV-2 in decomposed tissues. A few studies show that in aerosolized form, the virus can survive at relative humidity (RH) of 40-60% at a temperature of 19-22 °C for 90 mins, which is twice as longer as Influenza virus[27],[28]. The virus inoculated on various surfaces survived up to 28 days at 20 °C at 50% RH[29], pointing to the environmental stability of the SARS-CoV-2 virus under favourable temperature and humid conditions. Further, studies done on similar enveloped viruses such as influenza may also provide indication on the stability of SARS-CoV-2 in decomposed tissue. It was found that 90% of the virus gets inactivated in around fifteen days in the muscle, ten days in feather, and less than a day in the liver tissue when the carcass of the dead bird was left at room temperature. But if the carcass was preserved at refrigeration (4°C), the viability of the virus lasted 4.5 times longer, that is, more than two months[30]. Therefore, the stability of the virus in dead tissues depends on the localization of the virus in tissues at the time of death, temperature, humidity, number of copies of the virus, etc. In general, virus survival in dead tissue may not be longer as microbes will start to decompose the body and produce heat, which will limit the virus viability further. Considering the presence of cool, humid and moist location surrounded by the paddy fields, the presence of virus genome might also indicate the virus survival in decomposed tissues. However, culturing of the virus from swab or tissue samples and subsequent detection of viral proteins could confirm the virus viability in tissues after death.

Our findings also reveal implications to spatial and temporal distribution of SARS-CoV-2 in tissues at the time of death. Studies have shown that SARS-CoV-2 localizes in the lung and in other tissues including the kidney during advances stages of the disease[31]. However, it is not clear the factors that determine the spread of the virus to extrapulmonary tissues. Several studies correlate to higher expression levels of angiotensin converting enzyme (ACE)-2 receptors in some organs than in others[32]. Similarly, the presence of virus genome in the decomposed intestine and the liver in our study might correlate with higher expression of ACE2 receptors in them however, the absence of virus genome in the kidney and the lung might be attributed to the early infection stage. Further, subject had died without any apparent COVID-19 associated complications two days post-detection of the virus genome in naso-oropharyngeal sample by RT-PCR. Therefore, it is possible that at the time of death virus infection might be in the early stages with lesser viral load and without respiratory distress. Following swallowing of sputum, infection might have spread to the intestine that has substantial number of ACE-2 receptors, then to the liver through portal circulation. Further studies such as virus culture, detection of virus titre, and expression levels of other viral proteins are required to evaluate the virus survival in these samples.

In summary, we demonstrate for the first time that SARS-CoV-2 was detected in autolysed naso-oropharyngeal swabs and in tissues from an exhumed decomposed body. This report signifies the genome stability and also implicates survival stability of the SARS-CoV-2 in decomposed tissues, and swabbing upper respiratory mucosa is sufficient for obtaining samples for diagnosis. Further, results suggest the spatial distribution of the virus in tissues during early stage of infection with no respiratory distress. Additionally, findings also suggest that SARS-CoV-2, similar to other viruses such as Ebola virus[24], can persist for more than seven days on the surface of dead bodies, confirming that transmission from deceased subjects is possible for an extended period after death. These results further reaffirm the robustness of the RT-PCR aiding in the detection of viruses or their genome in exhumed, decomposed samples when other methods of detection could not be useful.

## Data Sharing

We declare we no objection to sharing and dissemination of this research findings.

## Data Availability

We declare we no objection to sharing and dissemination of this research findings.

## Acknowledgements

This study was supported by the financial support from the CSIR-CFTRI, Bharatiya Reserve Bank Note Mudran (P) Limited, Mysuru and Govt. of Karnataka. The corresponding author acknowledges, Department of Biotechnology (DBT), Govt. of India, for providing the Ramalingaswami Fellowship. All authors are thankful to the Director, CSIR-CFTRI for providing facilities to carry out this study.

